# Mass-Standardised Quantitative Measurements of the Antibody Levels for SARS-CoV-2 beyond Correlates of Protection and Clearance

**DOI:** 10.1101/2022.07.12.22277533

**Authors:** Philip H. James-Pemberton, Mark W. Helliwell, Rouslan V. Olkhov, Shivali Kohli, Aaron C. Westlake, Benjamin M. Farrar, Andrew M. Shaw

## Abstract

A quantitative biophotonic assay for IgG antibodies for the SARS-CoV-2 spike protein has been standardised against the NISTmAb antibody. Serum from patients recovering from infection, a sterilising serum, and post-vaccination, are used to profile anti-spike protein IgG responses. The vaccine response distributions were used to quantify live-challenge protective thresholds: the 31^st^ percentile of the AstraZeneca ChAdOx1-S (AZ, *n* =35 patients) distribution (2.90 ± 1.10 mg/L) and the 7^th^ percentile of the Pfizer/BioNTech BNT162b2 (Pfizer, *n* =25 patients) distribution (1.11 ± 1.10 mg/L). The recovery serum antibody response was characterised in 195 SARS-CoV-2 RT-PCR-positive patient and compared with 200 pre-pandemic patient samples. The diagnostic cut-off for RT-PCR-positive recovering patient samples was 1.34 ± 1.10 mg/L. The Anti-SARS-CoV-2 NISBC Verification Panel showed a separation of seropositive and seronegative samples at 1.90 ± 1.10 mg/L. The mean mass-standardised value of the two prevention and two recovery thresholds is 1.8 mg/L (95% CI 0.2–3.4) mg/L: moving beyond correlates of protection and clearance, to mechanism.

**Funding:** The work was funded by donations to Prof Shaw’s research group from the Exeter University Alumni during the pandemic, by Attomarker Ltd that funded PhD studentship for Philip James-Pemberton at the University of Exeter and Attomarker Ltd directly.

**Declaration of Interest:** Prof Shaw is the Founder, CEO and Director of Attomarker Ltd, a spin-out company from his research group.

**Related Publications in the Series:** **Paper 1** – This Paper - Mass-Standardised Quantitative Measurements of the Antibody Levels for SARS-CoV-2 beyond Correlates of Protection and Clearance

**Paper 2** - Mass-Standardised Differential Antibody Binding to a Spectrum of SARS-CoV-2 Variant Spike Proteins: Wuhan, Alpha, Beta, Gamma, Delta, Omicron BA.1, BA.4/5, BA.2.75 and BA.2.12.1 variants - Antibody Immunity Endotypes

**Paper 3** –Mass-Standardised Antibody Affinity Maturation to the Spike Protein of SARS-CoV-2 Omicron Variants in a Constant-Exposure Cohort: Forgiving Original Antigenic Sin

**Paper 4** – Diagnostic Classification for Long Covid Patients identifying Persistent Virus and Hyperimmune Pathophysiologies leading to a Test-and-Treat Protocol

## Introduction

Vaccine development, licensure and use would be significantly accelerated by the availability of harmonised correlates of protection^1^ (CoP); better still by mechanisms of protection. However, CoPs remain poorly determined^2-6^ in part because of the variability between neutralising antibody assays^7^ which prevent direct comparisons between cohorts and laboratories. Cohort comparison or immuno-bridging between diseases or variants, in the case of SARS-CoV-2 vaccines, would be ideal to allow prediction of vaccine protection against evolving variants. CoPs from 26 vaccines have been explored previously^4-6,8^ reporting results in a variety of arbitrary units with only three presented in mass units. These data point to a phenomenological protective serum which is currently not characterised and, consequently, with no disease-dependent understanding. Similarly, complete recovery from infection and removal of the virus from the body requires a further characterisation of a sterilising serum, preventing viral persistence and consequent post viral sequalae^9^ such as long covid.

Reproducible serum characterisation requires harmonisation across cellular neutralising antibody (NAb) assays each with different pseudovirus particles, receptor surfaces, signal transduction pathways and incubation times leading to non-standardised antibody titres and difficulty in making inter-cohort, inter-laboratory comparisons^10^. Standard materials take time to develop; those derived for SARS-CoV-2 serology include WHO reference materials and NIBSC samples which both define an arbitrary unit 1000 U/mL. Alternatively, non-cellular assays can be fully standardised against a the NIST standard human antibody^11^ specific to the respiratory syncytial virus protein F (RSVF)^12^, which can be used uniquely to calibrate a biophotonic assay platforms and quantify mass. The mass standard can be used to calibrate a humanised antibody raised to the SARS-CoV-2 Spike (S) protein, which in turn can be used to characterise the patient antibody response quantitatively for protective and sterilising sera.

Selection of a ‘control’ cohort for any SARS-COV-2 study now (2025) is almost impossible (unless from a pre-2019 biobank) given the spectrum of vaccination, symptomatic and asymptomatic infections present in the population. However, in the initial naïve vaccination challenge studies, the Pfizer vaccine showed an efficacy of 93% for reducing symptomatic cases^13^, suggesting a vaccine antibody concentration CoP at the lower 7^th^ percentile of the distribution of antibody vaccine response.

Similarly, the AstraZeneca vaccine had an equivalent efficacy of 69% – 74% in a real-world setting^14^: the lower 31^st^ percentile potentially corresponds to a similarly protective immunity level. The Moderna vaccine, likewise, demonstrated a 95.2% efficacy (95% confidence limits 91.2 % – 97.4 %)^15^ against infection. However, protection is transient with antibody half-lives reported between 60 – 200 days for natural antibodies following infection^16^ by the initial Wuhan variant or vaccination. A poor response to the vaccine may only provide protection above the threshold for a few tens of days.

In this paper, we investigate antibody immunity thresholds in different patient cohorts using a mass-standardised quantitative immunoassay for SARS-CoV-2 S IgG antibodies, standardised against the mass of the NIST reference human antibody. The test is used to profile the anti-S IgG serum response in patients receiving the AstraZeneca (AZ) and Pfizer vaccines as immune-naïve patients at the beginning of the pandemic. Estimates of the IgG protection thresholds can then be derived from the vaccine response IgG distributions: vaccine efficacy at preventing infection following live challenge 31^st^ percentile of the AZ distribution and 7^th^ percentile of the Pfizer vaccine response curves. Further, the sterilising serum has been characterised following the recovery of 196 PCR(+) samples early in the pandemic to the circulating Wuhan variant only compared with 200 PCR(-) negative pre-pandemic samples. The test is further used to measure the separation between positive and negative NIBSC standard samples. The four threshold estimates are then averaged to produce SARS-CoV-2 mechanistic threshold which is discussed in the context of mucosal immunity mechanisms of protection and viral clearance.

## Results

The mass-standardisation of the biophotonic assay to Spike S2 region produces a fully quantitative estimate of the response profiles of the patients to the Pfizer and AZ vaccines, Figure 1. Threshold levels associated with the lower 7^th^ percentile of the Pfizer distribution are plotted to correspond with the 93% live-challenge efficacy at protecting against infection; similarly, the 31^st^ percentile for AZ is shown, plotted to correspond with 69% efficacy. The commercial clinical sample threshold and the NIBSC sample threshold are also indicated in Figure 1, as are the three patients who received Pfizer booster doses, Table 1.

**Table 1.** Summary of immunity thresholds for vaccine responses, including preliminary literature survey (Error bars are standard deviations from nearest calibration points)

| Threshold | Cohort | Comments |
| --- | --- | --- |
| 1.34 ± 1.10 mg/L | Recovery Cohort Clinical threshold | 196 (+) samples<br>200 (-) samples<br>85% Sensitive<br>93% Specificity |
| 2.90 ± 1.10 mg/L | AstraZeneca: 31<sup>st</sup> percentile | 35 patient clinic samples (27 samples above LoD used) |
| 1.11 ± 1.10 mg/L | Pfizer: 7<sup>th</sup> percentile | 25 patient clinic samples (21 samples above LoD used) |
| 1.90 ± 1.10 mg/L | Youden threshold NIBSC Anti-SARS-CoV-2 Verification Panel | 22 (+) samples<br>14 (-) samples<br>82% Sensitivity<br>100% Specificity |
| 1.8 (95% CI 0.2 – 3.4) mg/L | SARS-CoV-2 Infection Immunity Threshold |  |
| 0.35 mg/L | *Pneumococcus* | for children<sup>4</sup> 7-component vaccine |
| 1 mg/L | *H. influenzae* | long-term protection<sup>20</sup> |
| 1 mg/L | *Haemophilus influenzae* | type b (Hib), Hib polysaccharides<sup>8,4</sup> |
| 5 mg/L | *H. influenzae* | Preventing colonisation of the nasal pharyngeal cavity<sup>4</sup> |

**Table 2.**
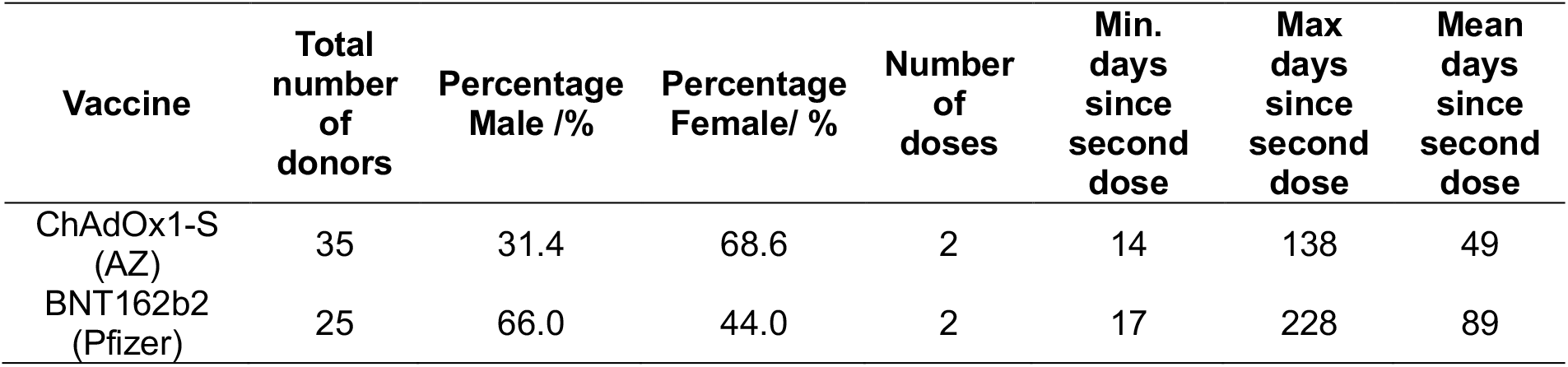
Demographic data for samples from double vaccinated individuals collected in the Attomarker clinic.

| Vaccine | Total number of donors | Percentage Male /% | Percentage Female/ % | Number of doses | Min. days since second dose | Max days since second dose | Mean days since second dose |
| --- | --- | --- | --- | --- | --- | --- | --- |
| ChAdOx1-S (AZ) | 35 | 31.4 | 68.6 | 2 | 14 | 138 | 49 |
| BNT162b2 (Pfizer) | 25 | 66.0 | 44.0 | 2 | 17 | 228 | 89 |

**Table 3.** Demographic data for samples collected and tested by Attomarker where the patients had received a Pfizer booster vaccination.

| Patient | Gender | Age | Days since Booster |
| --- | --- | --- | --- |
| A | Female | 25 - 34 | 37 |
| B | Male | 25 - 34 | 17 |
| C | Male | 75+ | >59 |

**Figure 1.**
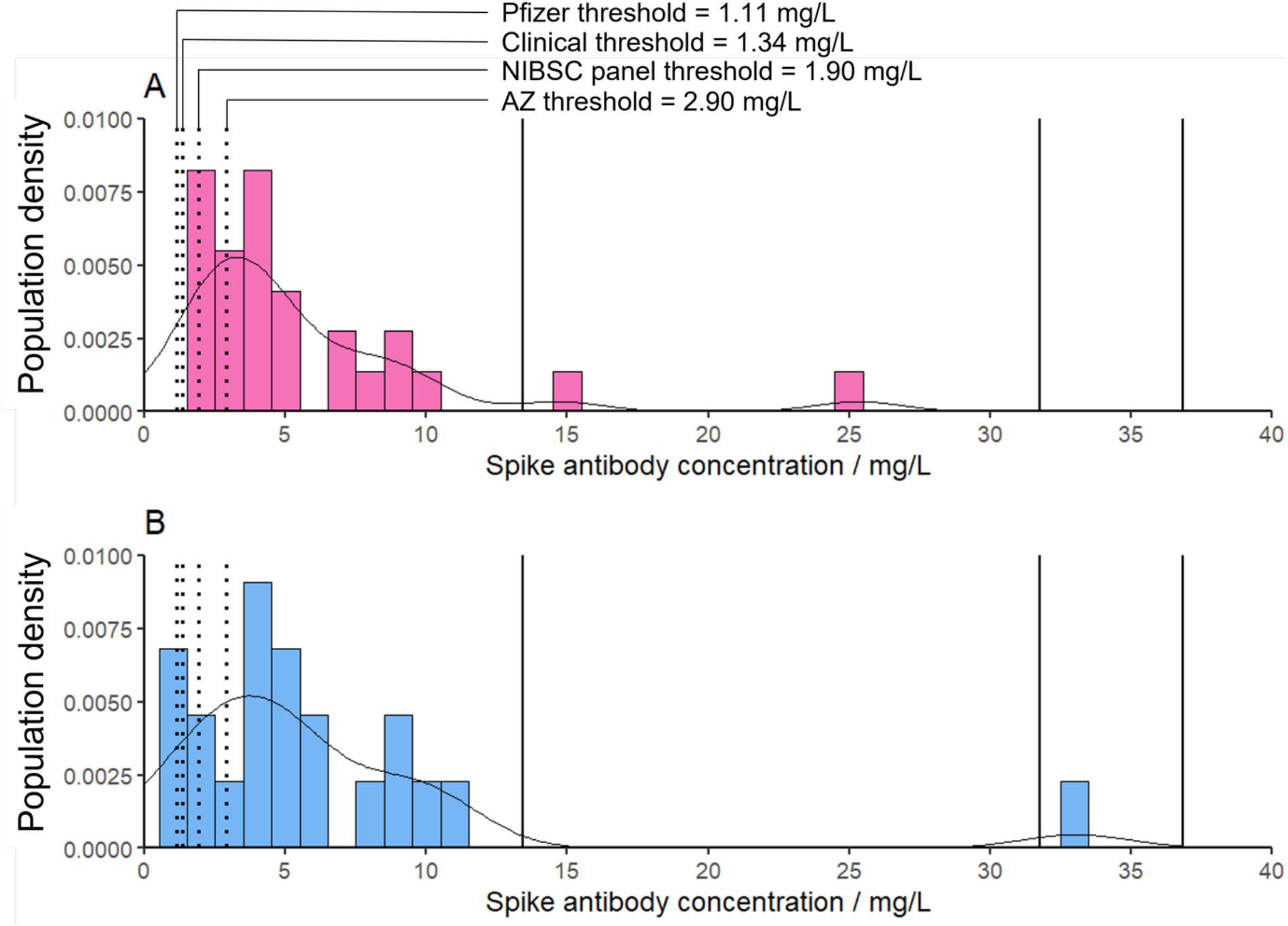
The fully quantitative spectrum of vaccine responses, showing all four immunity thresholds plotted on each graph: (A) the AZ response spectrum; and (B) the Pfizer response spectrum. The vertical lines (solid black) are individual patients who had received a booster vaccination.

The AZ IgG response distribution ranges from 0.6 mg/L – 25.4 mg/L with a distribution mode of 3.3 ± 1.0 mg/L whereas the Pfizer response distribution ranges from 0.6 mg/L – 33.1 mg/L with a distribution mode of 3.7 ± 1.0 mg/L. These data suggest that the immune response to the vaccine is highly personal. However, some of the ‘super responders’ had positive SARS-CoV-2 RT-PCR tests before vaccination, indicating the vaccine augmented the natural response.

The NIBSC sample responses are shown in Figure S4, along with their calibrated values. These data have been plotted against four other anti-SARS-CoV-2 serology technique results from the same samples, Figure 2A-D, with correlation coefficients of 0.77, 0.72, 0.51 and 0.62, respectively. The manufacturer conversion factors are shown in Table S1.

**Figure 2.**
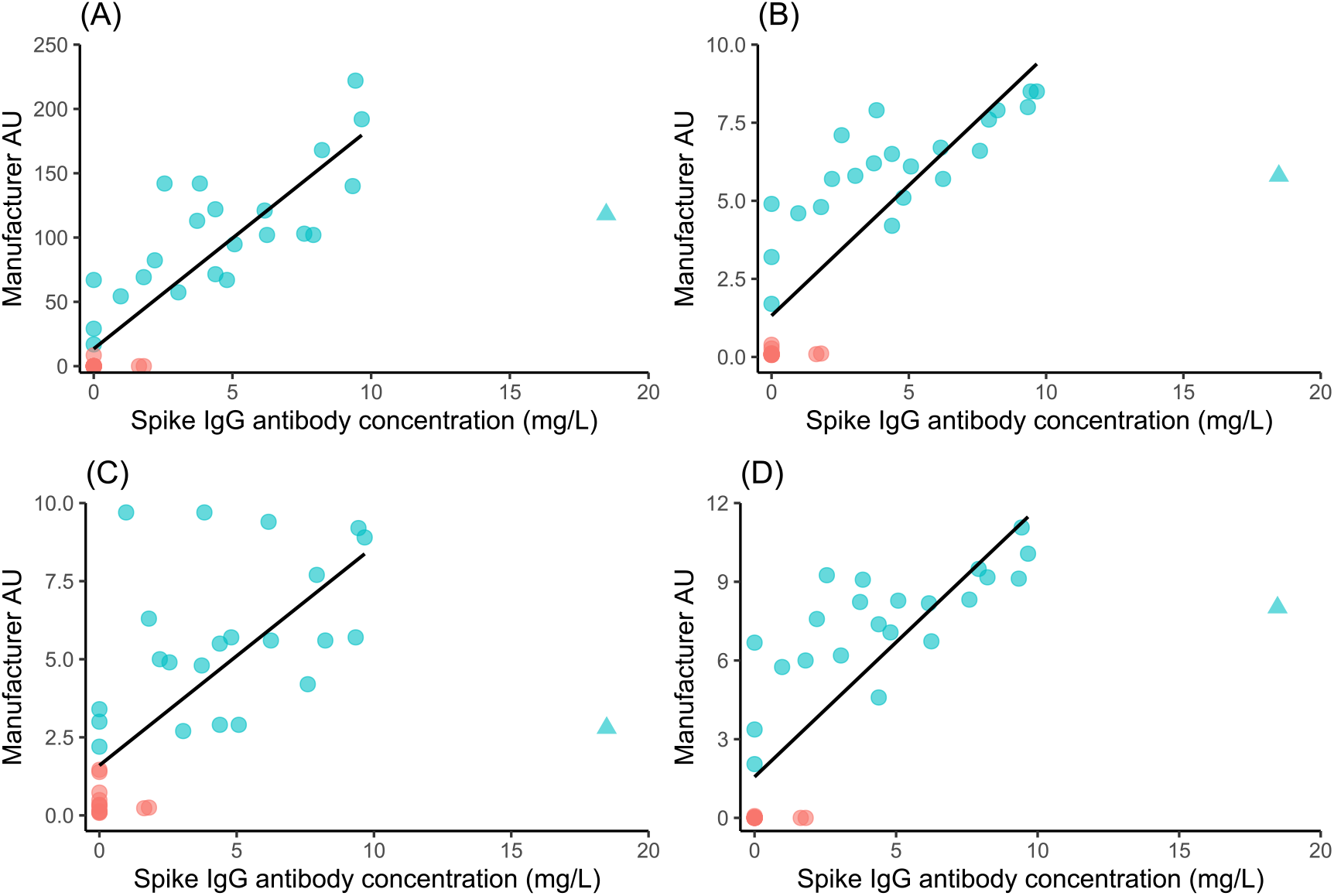
Plots of quantified Spike IgG Antibody result in mg/L vs four external tests for the NIBSC 20/B770 sample panel. Test platform (A) measures S1/S2 IgG; platform B ELISA S1 (recombinant) IgG; platform C ELISA S1 IgA and platform D S1 IgG. Sample with NIBSC sample ID 19 has been excluded from the fit. Pink circles are NIBSC negatives samples; Blue circles are positive NIBSC samples. The point depicted as a triangle is an outlier despite several repeats and was removed from the fit.

## Discussion

SARS-CoV-2 antibody testing, and more generally, all serology testing, is not well standardised with reports of 100-fold variation in standard sample measurements between laboratories^17^. Further, results are reported in variety of in-house, arbitrary units and more widely as antibody titre. The mass-standardised biophotonic assay is fully calibrated and can be readily certified. It is, however, biased towards antibodies that can bind to the target protein in under 2 minutes and remain stable during a 1-minute washing step and subsequent 2-minute detection step. The high-association rate antibodies form a likely subset of antibodies detected in other NAb assays because of varying incubation times. Further, the spike antibody assay is restricted to anti-spike IgG and does not include other membrane or envelope protein antibodies^18^ which may also be neutralising (preventing endocytosis for example). However, the biphotonic binding assay affinity selection, typically nM or better, reflects the affinity maturation of antibody production within the body^19^.

The biophotonic assay platform is intrinsically mass sensitive and therefore a linear response with mass. The correlation with other techniques is highly variable, as seen in Figure 2, with correlation coefficients varying between 0.51 - 0.77, with some techniques being intrinsically non-linear (Figure 2B - 2D). There is also non-standardisation in the detection targets, some choosing IgG only, whilst others include IgM and IgA. The non-standardisation makes comparison between cohorts very difficult, leading to laboratory-specific characterisation of vaccine responses and immunotherapy. Fundamentally, this impacts the development, use and licensure of vaccines.

The mass-standardised approach reported here presenting results in mg/L, allows the absolute characterisation of sterilising and protective sera. The two vaccine response distributions identify quantitatively the lower 7^th^ percentile for Pfizer^13^ and 31^st^ percentile for AZ^14^, the protective thresholds in the live-virus challenge studies, Table 1, which are similar to the recovery thresholds derived from the PCR(+/-) cohort. Likewise, the standardised NIBSC sample cohort are near-coincident when measured quantitatively. The average of all four thresholds investigated in this study suggests a mean of 1.8 mg/L, with 95% confidence limits of 0.2 – 3.4 mg/L which would represent the threshold for all variants if the mechanism of protection is prevention of colonisation in the nasal mucosa. The vaccine distributions also show that protection is transient with ∼40% of the population within one half-life of the threshold, similarly, the potential for incomplete recovery leading to persistent virus. The antibody half-life variation suggests a boosting schedule that should be personalised.

Mass standardisation allows CoPs to be replaced with mechanisms of protection or clearance in recovery. Nasal IgG levels, secreted from the blood, have been shown to prevent nasopharyngeal colonisation by *H. influenzae* with specific IgG levels of 5 mg/L or higher in the plasma^21^. Neutralisation of SARS-CoV-2 virus particles requires a proportion of the spike proteins to be bound to antibodies preventing binding to the ACE2 receptors during the residence time of the complex. Whilst the nasal mucosa contains a T cell density of 650 mm^-2^ (with a factor of 5 variation)^22^ which would sufficiently remove infected cells, there is also total IgG^23^ 4.5 – 44 (median 16.4) ×10^13^ IgG molecules per mL, of which 10^11^ would prevent the virus particle binding to the ACE2 receptor (as predicted by the thresholds derived here). Assuming that the fraction of total IgG which is anti-Spike is the same in the nasal mucosa as in the serum, 1.8 mg/L corresponds to 10^11^ antibodies per mL. A challenge of 10^9^ viral particles^24^ would be complexed by this concentration, preventing ACE2 receptor binding, with any breakthrough infections due to overwhelming large viral loads.

Different parts of the population may have different thresholds depending on ACE2 receptor distributions and mucus production, which also points to differences between respiratory diseases and their target receptor distributions. Similarly, the IgG serum threshold also represents sufficient antibody concentrations in the blood to distribute to all body volumes and clear the virus, resulting in a complete recovery.

The protective concentration of 1.8 mg/L, 0.2 – 3.4 mg/L (95% CI) should be variant independent as long as there are sufficient antibodies to complex the virus. Insufficient antibodies will lead to vaccine breakthrough. Uniquely in the cohorts here, this was 7% for the Pfizer vaccine and 31% for the AZ vaccine, specifically for one variant where all antibodies have identified active epitopes on the target spike protein. Comparatively small changes in antibody concentrations would lead to infection or increased viral load. By comparison, CoPs are variant specific^25^. Five-fold variation Receptor Binding Domain (RBD)-ACE2 binding affinities have been observed for different variants^26^; five-fold RBD-ACE2 association and dissociation rates^27^ also show variant dependence; and more interestingly increased viral load in later variants^28^. However, the mechanism of antibody binding preventing the encounter of the 90 nm viral particle with the ACE2 receptor nasal distribution requires a similar number of antibodies: if a mutation in the spike protein renders only 50% of the target epitopes active in the IgG distribution, then a patient will have a variant-dependent breakthrough immunotype, which includes all parts of the immune system response.

Protection and clearance concentrations are however transient, with natural antibody half-lives varying 60 – 200 days following infection^16^. Similarly, if the antibody quality at the pH of the relevant mucosa is not sufficient (nM affinity) then the prevention or clearance will be compromised; high affinity antibody-virus complexes will have a half-life of hours. Poorly matured antibodies or variations in the effective epitope spectrum could lead the NAbs in a proportion of the population becoming ineffective against a specific variant in nasal protection and vaccine, leading to breakthrough infections or persistent virus in recovery. These mass-standardised measures are critical to vaccine design and deployment for the management of future pandemics, for example, the delivery programmes to eliminate malaria^29,30^. Mass-standardisation would also improve our general understanding of immunology and vaccine development.

## Methods and Materials

### Methods

#### Biophotonic Multiplexed Immuno-kinetic assay

The core technology is an immuno-kinetic assay using a biophotonic detection event and has been described in detail elsewhere in a SARS-CoV-2 antibody sensing application^18^. Briefly, an array of gold nanoparticles is illuminated in a total-internal-reflection configuration exciting a localised plasmon in the particles scattering the light. The intensity of scattered light depends on the mass in the plasmon field (formally permittivity) so a greater mass of proteins leads to greater intensity in scattered light from the array surface. This signal is captured as a video image in real time. The multiplexed nanoparticle array is functionalised with capture molecules to give analytic specificity to the target analyte. The array of the Attomarker CE marked COVID-19 Antibody Immunity Test consists of five tests: total IgG captured by Protein A/G (PAG), polyclonal goat antibodies to measure C-reactive protein (CRP), and the SARS-CoV-2 proteins Nucleocapsid (N), Spike (S) and Receptor Binding Domain (RBD). In addition, a recombinant human serum albumin (HSA) channel acts as a control to adjust for temperature at the array, non-specific binding and light intensity variation. A 20 *μ*L serum sample is diluted 100-fold, centrifuged and then fat-filtered, before being flowed over the array. The brightness change in the scattered light intensity is integrated over 120 s during a capture step, which is followed by a wash step to remove unbound protein and low affinity antibodies, followed by a quantitative detection step using anti-analyte antibodies (integrating for 90 s), completing a sandwich assay to deliver results in 7 minutes.

The response of the biophotonic array is calibrated using the international NIST standard which is a high-purity, monomeric, recombinant IgG1k with a known sequence^12,31^. The purity of the antibody is measured using the PAG assay on the array, which binds antibodies specifically via the Fc region. The Antibody Immunity Test can then be calibrated in a two-step process: 1) Calibration of the site density of the PAG channel using a 7-point calibration curve derived from the NISTmAb; and 2) Determination of the concentration of a humanised mouse antibody specific to the SARS-CoV-2 Spike protein S2 subdomain and the subdomain integrity (Figure S1). The Spike antigen site density is then calibrated using three concentrations using the humanised mouse antibody to span the concentration range in the samples.

The PAG assay was calibrated over an extended calibration curve of 1.56 – 50.0 nM using the NISTmAb and the Spike protein assay was linear over the range 0.6 – 20.0 mg/L (Figure S2). All Antibody Immunity Test sensor chips were calibrated with 3-point calibration using either 1.0, 5.0 and 20.0 mg/L (100-fold dilution) or 1.0, 5.0 and 10.0 mg/L (50-fold dilution) calibrant samples, each measured twice. The experiments were repeated with 100-fold and 50-fold dilutions to establish dilution invariance, with a limit of detection of 1.6 mg/L for 100-fold dilution and 0.6 mg/L for 50-fold dilution. A set of NIBSC samples was also measured to allow comparison between techniques. The threshold for the Antibody Immunity Test was calibrated using two commercial samples close to the threshold and determined by maximising the Youden Index^32^.

Four immunity thresholds were then calculated from five patient data sets. The antibody responses of double vaccinated patients (AZ *n* = 35, Pfizer *n* = 25) were measured with full consent. The experimental response distributions were plotted, from which percentile concentrations for vaccine efficacy were derived based on figures reported in Phase III clinical trials of both vaccines (AZ 69%; Pfizer 93%). Further immunity thresholds were determined using the separation between positive and negative NIBSC samples and the clinical threshold for the Antibody Immunity Test. The error estimates for the four immunity thresholds were determined from the repeatability of calibration samples at similar concentrations.

### Materials

Materials used throughout the course of the experiments were used as supplied by the manufacturer, without further purification. Sigma-Aldrich supplied phosphate buffered saline (PBS) in tablet form (Sigma, P4417), phosphoric acid solution (85 ± 1 wt. % in water, Sigma, 345245) and Tween 20 (Sigma, P1379). Glycine (analytical grade, G/0800/48) was provided by Fisher Scientific. Assay running and dilution buffer was PBS with 0.005 v/v % Tween 20 and the regeneration buffer was 0.1 M phosphoric acid with 0.02 M glycine solution in deionized water.

The recombinant Human Antibody to the Spike protein S2 subdomain was a chimeric monoclonal antibody (SinoBiological, 40590-D001, Lot HA14AP2901). The antibody was raised against the following immunogen: recombinant SARS-CoV-2 / 2019-nCoV Spike S2 ECD protein (SinoBiological, 40590-V08B). C-reactive protein (CRP)-depleted serum from BBI solutions (SF100-2). NISTmAb, Humanized IgG1κ Monoclonal Antibody from National Institute of Standards and Technology (RM8671). The NISTmAb is a recombinant humanized IgG1k with a known sequence^31^ specific to the respiratory syncytial virus protein F (RSVF)^12^. The detection mixture consisted of a 200-fold dilution of IG8044 R2 from Randox in assay running buffer.

COVID-19 Triple Antibody Test sensor chips were printed with recombinant human serum albumin from Sigma-Aldrich (A9731), casein from Thermo Scientific (37582), anti-CRP from Biorad (1707-0189G), recombinant Protein A/G from Thermo Scientific (21186), SARS-CoV-2 Spike Protein (RBD, His tag) from GenScript (Z03479-100), SARS-CoV-2 Spike S1+S2 ECD-His Recombinant protein from SinoBiological (40589-V08B1) and SARS-CoV-2 Nucleocapsid protein from the Native Antigen Company (REC31851-100).

### Patient Samples

#### 400 commercial samples

A set of 400 serum samples was purchased from two suppliers (Biomex GmbH and AbBaltis) to derive the sensitivity, specificity and clinical threshold for the Triple Antibody Test. Negative samples: 200 pre-pandemic (pre-December 2019) human serum samples were purchased from AbBaltis. All were tested and found negative for STS, HBsAg, HIV1 Ag(or HIV PCR(NAT)), HIV1/2 antibody, HCV antibody and HCV PCR(NAT) by FDA approved tests; and positive samples: 90 samples were purchased from AbBaltis which were all from PCR+ individuals. They were also tested for IgG and IgM antibodies to SARS-CoV-2; all 90 were positive for IgM antibodies and 70 were positive for IgG antibodies. No information was provided regarding symptoms of the donors. 45% of these samples were from Female donors and 55% were from Male donors. The age of donors ranged from 19-81 years. No information on time from infection to sample collection was given.

106 samples purchased from Biomex were from PCR+ individuals. All samples were YHLO Biotech SARS-CoV-2 IgG positive and Abbott SARS-CoV-2 IgG positive. A spectrum of symptoms from the following list were detailed for each sample: fever, limb pain, muscle pain, headache, shivers, catarrh, anosmia, ache when swallowing, diarrhoea, breathing difficulties, coughing, tiredness, sinusitis, pneumonia, sickness, lymph node swelling, pressure on chest, flu-like symptoms, blood circulation problems, sweating, dizziness and hospitalisation. Time from infection to sample collection ranged from 27 – 91 days. 35% of these samples were from Female donors and 65% were from Male donors.

The commercial sample set was tested with the Attomarker Qualitative Triple Antibody test (CE Marked) without removing any samples. The test uses Boolean logic classifying samples as positive if Nucleocapsid, Spike or Receptor Binding Domain above a threshold. The sample showed a sensitivity of 92%, specificity of 91% and unclassified of 10%, including 1 triple false positive and two double false positives, which are likely to be miss-classifications. The Boolean thresholds were calibrated in the current study quantitatively by remeasuring samples close to the cut-off.

#### Clinical samples

Samples were collected from patients in Attomarker partner clinics, all of whom provided informed consent for their anonymised data to be used in research to aid the pandemic response. The data from tests of 63 patient samples are included in this study. 60 had received two doses of either the AstraZeneca SARS-CoV-2 vaccine or the Pfizer SARS-CoV-2 vaccine at least 14 days prior to sample collection and testing by Attomarker. Three further samples had received a third vaccination.

#### National Institute of Biological Standards and Control (NIBSC) SARS-CoV-2 Standard samples

The NIBSC Anti-SARS-CoV-2 Verification Panel for Serology Assays (product code: 20/B770) was purchased. This panel contains 37 samples. 23 samples are from convalescent plasma packs which are known to be Anti-SARS-CoV-2 positive; 14 samples are plasma packs known to be Anti-SARS-CoV-2 negative.

## Data Availability

All data produced in the present study are available upon reasonable request to the authors.

## Ethics

The samples were collected with informed patient consent for use in better understanding pandemic. The use of the samples has been reviewed independently by the Biosciences Ethics Committee, University of Exeter and approved.

## Acknowledgements

The authors would like to thank Dr Jonathan Snicker and Mr Sydney Nash for their guidance on the potential strategic and policy implications of the scientific findings.

## Supplementary Data

**Figure S1.**
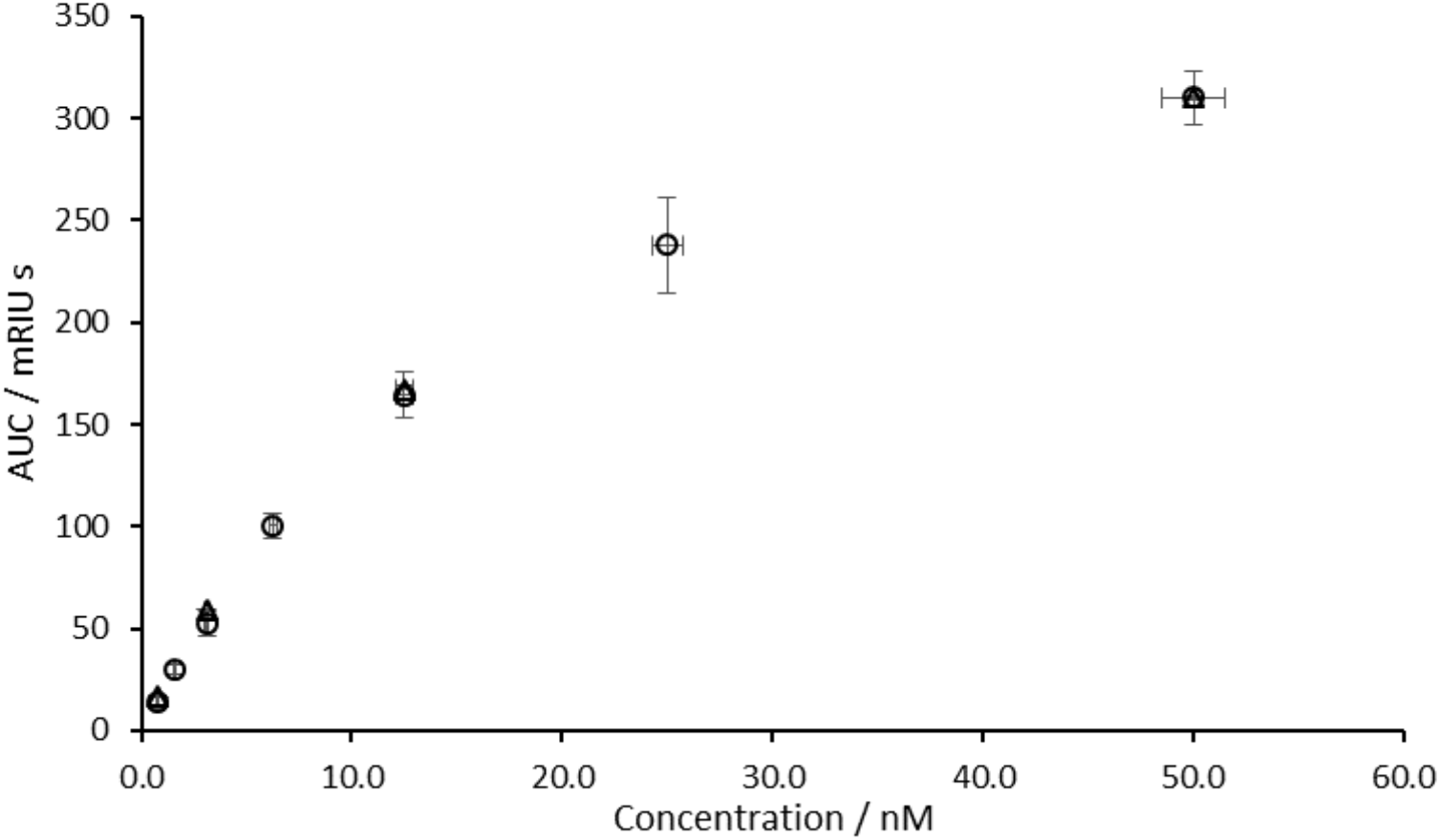
Measurement of the concentration and monomeric purity of the chimeric anti-Spike antibody (Sinobiological, 40590-D001) relative to the NIST supplied monoclonal standard RM8671. The response shown is the area-under-curve (mili Refractive Index Units seconds mRIUs) of the Protein A/G - rHSA channels. (○) denote samples of NIST monoclonal at concentrations of 50.0, 25.0, 12.5, 6.25, 3.13 and 1.56 nM. (△) denote samples of anti-Spike antibody at concentrations of 50.0, 12.5, 3.13 and 1.56 nM. Vertical error bars show 1 s.d. of repeats. Horizontal error bars show 3% error as the assumed possible error from sample pipetting.

**Figure S2.**
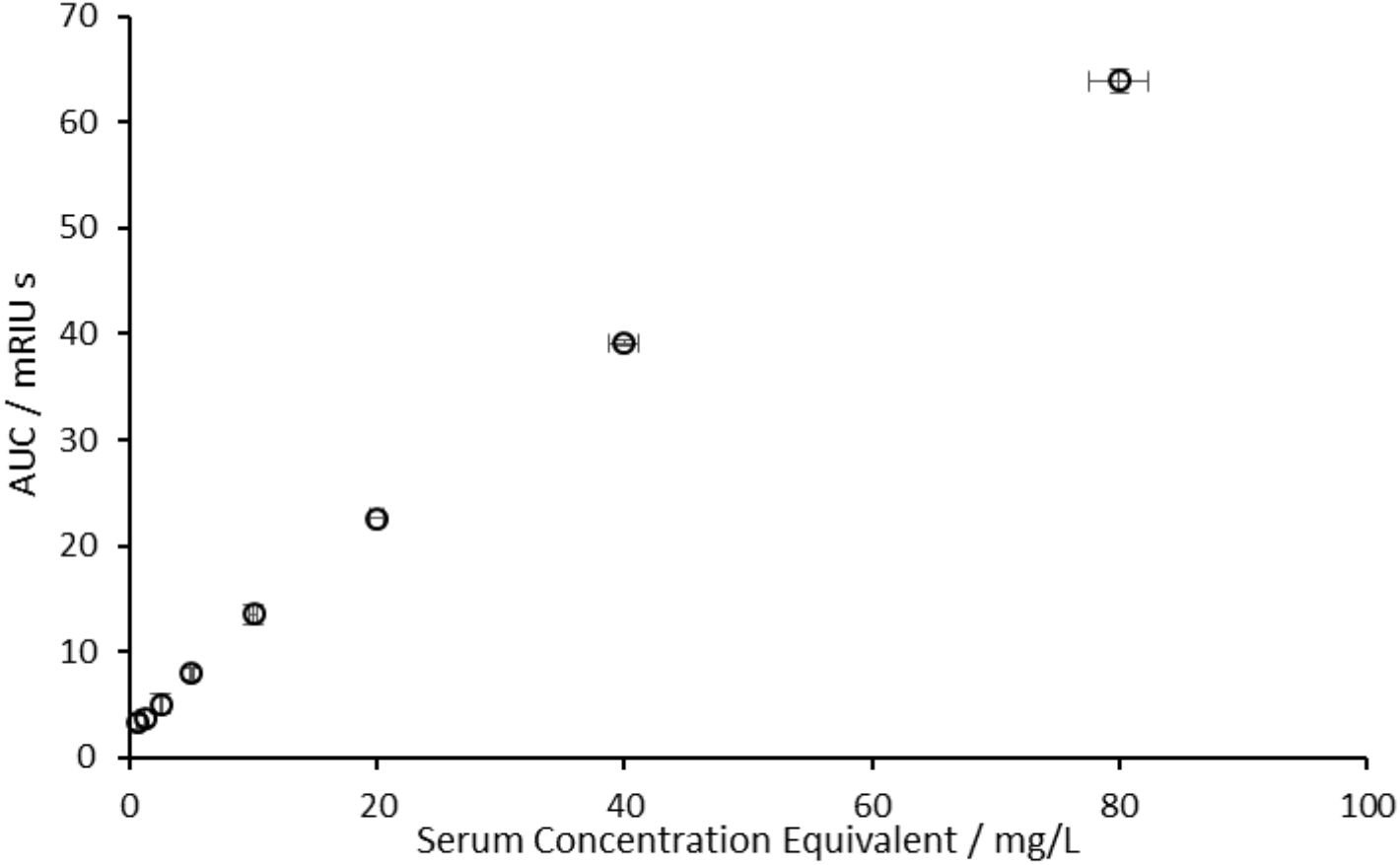
The response curve to samples of the chimeric anti-Spike antibody (Sinobiological, 40590-D001), flowed over the sensor for 120s each. The response shown is the area-under-curve of the Spike - rHSA channels. A detection mixture of 200-fold diluted Randox antiHuman IgG (IG8044) in PBS-T buffer was flowed over the sensor for 90s. Samples were made up in 100-fold diluted serum, obtained from BBI pre-pandemic (SF100-2), at concentrations of 5.33, 2.67, 1.33, 0.66, 0.33, 0.17, 0.08, 0.04 nM, corresponding to pre-dilution serum antibody concentrations of 80.0, 40.0, 20.0, 10.0, 5.0, 2.5, 1.3, 0.6 mg/L. Vertical error bars show 1 s.d. of repeats. Horizontal error bars show 3% error as the assumed possible error from sample pipetting.

**Figure S3.**
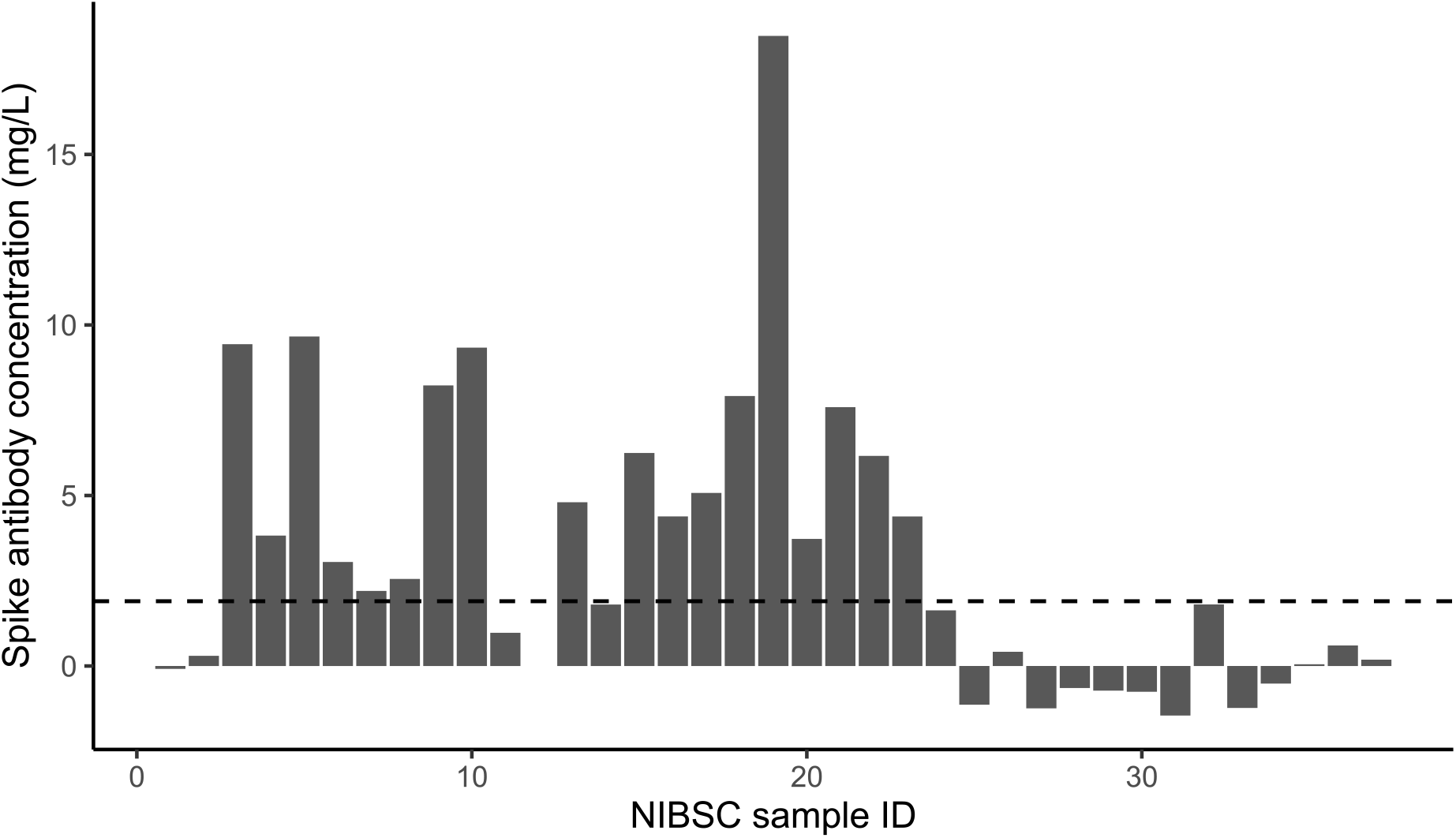
The calculated Spike antibody concentrations of a panel of NIBSC samples. Samples 1-23 are listed as positive; 24-37 are negative. Values were calculated using calibration samples of 40590-D001 at 1, 5, 10 and 20 mg/L. All sample measurements were single measurements, with calibrators repeated. Note that sample 12 has not been tested.

**Table S1.** Lines of best fit and Correlation coefficients for the comparison with four platform technologies against the current fully quantitative test.

| Platform | Slope | Intercept | R<sup>2</sup> |
| --- | --- | --- | --- |
| A | 17.2 | 13.5 | 0.77 |
| B | 0.83 | 1.32 | 0.72 |
| C | 0.70 | 1.60 | 0.51 |
| D | 1.03 | 1.56 | 0.69 |

